# Longitudinal symptom transitions predict incident tuberculosis risk in serial active case finding

**DOI:** 10.1101/2025.10.26.25338759

**Authors:** Esther Jung, José Victor Bortolotto Bampi, Alessandra Moura da Silva, Yiran E. Liu, Daniel Henrique Tsuha, Argita D. Salindri, Andrea da Silva Santos, Roberto Dias de Oliveira, Julio Croda, Jason R. Andrews

**Affiliations:** Stanford University School of Medicine, Stanford, California; Federal University of Mato Grosso do Sul, Campo Grande, Brazil; Yale University School of Medicine, New Haven, Connecticut; Yale Law School, New Haven, Connecticut; Federal University of Grande Dourados, Dourados, Brazil; State University of Mato Grosso do Sul, Dourados, Brazil; Oswaldo Cruz Foundation, Campo Grande, Brazil; Yale University School of Public Health, New Haven, Connecticut

**Author notes:** **Corresponding Author:** Jason Andrews, MD, SM, Stanford University School of Medicine, 240 Pasteur Dr, Stanford, CA 94304-1049. JC and JRA contributed equally to this manuscript.

**Keywords:** tuberculosis, asymptomatic, prisons, active case finding, screening

## Abstract

Tuberculosis (TB) screening often relies on cross-sectional symptom assessment. To determine the added value of longitudinal symptom monitoring, we conducted a prospective cohort study among 2,282 incarcerated men in Brazil. Every four months, participants were assessed for symptoms and incident TB (Xpert or culture positive). Compared to remaining asymptomatic, developing symptoms was associated with higher TB risk (aRR 2.31; 95% CI 1.55-3.43), strongest among those with prior radiographic abnormalities (aRR 2.49; 95% CI 1.56-3.97), while remaining symptomatic was not associated (aRR 1.27; 95% CI 0.72-2.24). Longitudinal symptom monitoring may enhance TB screening in high-risk settings, particularly in complement with radiography.

**GENERAL SUMMARY:** In an active case-finding program for TB, developing new symptoms was associated with over double the TB risk, while persistent symptoms were not. Monitoring symptom changes longitudinally, especially combined with chest radiography, may improve TB case detection in high-risk settings.

## BACKGROUND

Tuberculosis (TB) continues to pose a major global health challenge^1^. Early detection and treatment are essential to interrupt transmission and improve outcomes, particularly in high-risk populations. Efforts to identify individuals living with TB often leverage symptom screening to prioritize individuals for confirmatory molecular rapid diagnostic tests^2^. However, conventional approaches to TB screening typically only assess symptoms present at the time of screening entry, which may not capture the dynamic nature of symptom changes during TB disease progression^3,4^.

Symptoms that are typically classified as features of TB^5^, such as cough, fever, night sweats, and weight loss, can fluctuate over the course of disease^6^. In high-risk populations such as persons deprived of liberty (PDL), these symptoms may also be caused by other common conditions, including upper respiratory tract infections, pneumonia, chronic obstructive lung disease and tobacco smoking^7^. Moreover, recent studies reveal that over 50% of individuals living with undiagnosed TB do not report symptoms at the time of diagnosis^1,8^. Together, the non-specific presentation of TB and high prevalence of asymptomatic TB may limit the ability of cross-sectional symptom screening to reliably identify individuals at imminent risk of active TB, resulting in continued transmission^8,9^.

Longitudinal monitoring of symptoms offers an opportunity to overcome some of these challenges by capturing individuals’ changes in symptom status over time, including new onset, persistence, or resolution. This may provide additional predictive value for disease, improve identification of individuals at high risk of TB, or indicate higher risk for future evaluations, thereby facilitating the prioritization of confirmatory diagnostic testing.

Prisons harbor some of the highest TB incidence and transmission rates worldwide^10–12^, and the World Health Organization recommends systematic screening be performed among incarcerated populations at least yearly^2^. However, active case finding is not widely implemented in most prisons in high-burden countries, partly due to resource constraints. Understanding how TB symptoms evolve in such settings could inform more resource-efficient screening approaches, yet longitudinal symptom trajectories in high-risk populations remain uncharacterized.

To address this knowledge gap, we characterized longitudinal symptom trajectories in a cohort of incarcerated individuals who were evaluated for TB symptoms and tested for TB every four months. We assessed the relationship between symptom transitions and risk of incident TB.

## METHODS

We performed systematic screening for TB every four months from February 2023 to September 2025 in two all-male prisons in the Brazilian state of Mato Grosso do Sul. After providing informed consent to participate, participants completed a standardized symptom questionnaire, administered by study nurses, to ascertain the presence of cough, fever, night sweats, weight loss, chest pain, or dyspnea. Presence of each symptom at each visit was assessed through yes/no questions (i.e. “Do you have a cough?”). An individual was considered symptomatic if they confirmed the presence of any of these symptoms.

At baseline, defined as the first visit in which a participant was enrolled in the study, all consenting participants had a posterior-anterior chest radiograph performed and two sputum samples collected for testing by culture and Xpert^®^ MTB/RIF Ultra (hereinafter “Xpert”). Chest radiographs were evaluated using automated detection software LunitTB (South Korea; version v3.1.5.1), which outputs a numerical TB risk score on a continuous scale between 0 and 100.

At each four-month follow-up visit, procedures were repeated: symptom assessment, a single sputum sample collection, and chest radiography. Symptom status transitions over sequential four-month intervals were categorized as asymptomatic-to-symptomatic, symptomatic-to-asymptomatic, persistently symptomatic, or persistently asymptomatic. Incident TB was defined by at least one positive sputum culture or Xpert result. Trace Xpert results were repeated with a new sample. Those that persisted as traces were considered negative for TB and all trace samples were cultured.

Eligibility was restricted to those with negative baseline Xpert and culture results who did not initiate TB treatment, regardless of radiographic findings, to ensure participants did not have prevalent TB. However, during follow-up, a small number of microbiologically negative participants initiated TB treatment based on clinical judgement. Only participants who completed at least one follow-up visit during the study period were included, to permit longitudinal assessment of symptom status and incident TB.

Given the frequent transfers and movement within the prison system, nearly 25% of individuals enrolled in the cohort missed follow-up visits intermittently. We excluded individuals with non-consecutive symptom information, as gaps in reporting precluded reliable classification of symptom transitions across sequential four-month intervals into the predefined symptom trajectory categories.

We used the chi-square and Kruskal-Wallis tests to compare baseline demographic and clinical characteristics across symptom transition groups in the first four-month interval.

In supplementary analyses, we evaluated all four-month symptom transitions across the study period to identify predictors associated with developing or resolving symptoms. We performed modified Poisson regression with robust standard errors clustered at the individual level to estimate adjusted risk ratios (aRRs)^13^. Models adjusted for the following baseline covariates: age, prison, prior TB, exposure to symptomatic cellmates (yes/no), TB contact (ever/never), current smoking status, any drug use, duration of incarceration, and binary LunitTB x-ray score using a threshold of 50 as a proxy for lung abnormalities. Separate models were constructed for individuals who were asymptomatic at the prior visit (assessing the risk of becoming symptomatic) and for those who were symptomatic at the prior visit (assessing the risk of becoming asymptomatic).

Specific symptom-level transition patterns were summarized descriptively by calculating the proportion of each individual TB-related symptom reported at visits where a transition occurred, distinguishing between new onset, resolution, and persistent symptoms.

We also characterized how long participants remained in their initial symptom state during follow-up using survival analysis. For symptomatic individuals, the event was the first resolution of symptoms, while for asymptomatic individuals, the event was the first acquisition of symptoms. Participants were censored at TB diagnosis, loss to follow-up, or last study visit.

For the primary analysis, we assessed whether symptom transitions during the preceding four-month interval were associated with incident TB diagnosed at the subsequent visit, using the same modified Poisson framework described above but adjusting for radiographic status (Lunit score <50 vs. ≥50) at the start of each interval rather than at baseline. Results were also stratified by radiographic status. Persistently asymptomatic individuals served as the reference category. All analyses were conducted in R (version 4.4.1).

Ethical approval was obtained from the Research Ethics Committee of the Federal University of Mato Grosso do Sul, the Brazilian National Commission for Ethics in Research, and the Stanford University Research Ethics Committee.

## RESULTS

After applying inclusion criteria, our study identified 2,282 participants without TB at baseline who were assessed for symptoms at two or more consecutive visits between February 2023 and September 2025. The median baseline age of participants was 33 years (IQR 28-40), over 80% of participants were Black or mixed race (1,854/2,282), and most participants did not complete high school (83%) (Table 1).

**Table 1.** Characteristics of participants by symptom transition between baseline and first follow-up visit (N = 2,282)

| Characteristics | Overall<br>(N = 2,282) | First 4-month interval symptom transition |  |  |  | p-value* |
| --- | --- | --- | --- | --- | --- | --- |
|  |  | Persistently<br>asymptomatic<br>(n = 1,417) | Persistently<br>symptomatic<br>(n = 215) | Asymptomatic<br>to Symptomatic<br>(n = 330) | Symptomatic to<br>Asymptomatic<br>(n = 320) |  |
| Median age (IQR) | 33 (28, 40) | 33 (28, 40) | 33 (28, 40) | 32 (27, 39) | 33 (27, 40) | 0.208 |
| Incident TB | 50 (2.2) | 26 (1.8) | 4 (1.9) | 15 (4.5) | 5 (1.6) | 0.018 |
| Ethnicity |  |  |  |  |  | 0.204 |
| White | 419 (18.4) | 251 (17.7) | 40 (18.6) | 57 (17.3) | 71 (22.2) |  |
| Black | 217 (9.5) | 127 (8.9) | 27 (12.6) | 33 (10.0) | 30 (9.4) |  |
| Mixed | 1,637 (71.7) | 1,034 (73.0) | 145 (67.4) | 239 (72.4) | 219 (68.4) |  |
| Asian | 8 (0.4) | 4 (0.3) | 3 (1.4) | 1 (0.3) | 0 (0) |  |
| Indigenous | 1 (<0.1) | 1 (<0.1) | 0 (0) | 0 (0) | 0 (0) |  |
| Education |  |  |  |  |  | 0.562 |
| Less than high school | 1,893 (83.0) | 1,175 (82.9) | 181 (84.2) | 279 (84.5) | 258 (80.6) |  |
| Completed high school | 389 (17.0) | 242 (17.1) | 34 (15.8) | 51 (15.5) | 62 (19.4) |  |
| Prison |  |  |  |  |  | 0.256 |
| EPJFC | 1,745 (76.5) | 1,089 (76.9) | 165 (76.7) | 239 (72.4) | 252 (78.8) |  |
| IPCG | 537 (23.5) | 328 (23.1) | 50 (23.3) | 91 (27.8) | 68 (21.3) |  |
| Previous TB | 409 (17.9) | 225 (15.9) | 54 (25.1) | 67 (20.3) | 63 (19.7) | 0.004 |
| TB contact | 1,335 (58.5) | 818 (57.7) | 135 (62.8) | 204 (61.8) | 178 (55.6) | 0.239 |
| Symptomatic cellmates | 854 (37.4) | 415 (29.3) | 132 (61.4) | 126 (38.2) | 181 (56.6) | <0.001 |
| Current smoking | 1,342 (58.8) | 741 (52.3) | 165 (76.7) | 220 (66.7) | 216 (67.5) | <0.001 |
| Any drug use | 1,416 (62.1) | 829 (58.5) | 150 (69.8) | 226 (68.5) | 211 (65.9) | <0.001 |
| LunitTB Score at Baseline |  |  |  |  |  | 0.961 |
| <50 | 1,584 (69.4) | 984 (69.4) | 146 (67.9) | 230 (69.7) | 224 (70.0) |  |
| ≥50 | 698 (30.6) | 433 (30.6) | 69 (32.1) | 100 (30.3) | 96 (30.0) |  |
| Months incarcerated, median<br>(IQR) | 12 (2, 44) | 13 (2, 41) | 14 (2, 71) | 11 (2, 34) | 9 (1, 61) | 0.213 |

No (% of column n) unless otherwise specified.
\*Pearson's Chi-squared test; Kruskal-Wallis test

The majority of individuals reported no TB symptoms at baseline (1,747/2,282, 76.5%) and 62.1% (1,417/2,282) remained asymptomatic at the first follow-up. Notably, 52% (26/50) of incident TB cases occurred among participants who remained asymptomatic. Among those asymptomatic at baseline, 14.5% developed symptoms by the first follow-up. Of the 23.4% symptomatic at baseline, 40% remained symptomatic and 60% reported resolution (Table 1). Several characteristics differed significantly across transition groups, with individuals with persistent symptoms showing the highest prevalence of these factors: previous TB, exposure to symptomatic cellmates, current smoking, and any drug use (Table 1).

Of 2,282 total participants, 1,725 (75.6%) were assessed for at least 3 visits, 1,105 (48.4%) for at least 5, and 578 (25.3%) completed all 7 possible assessments. Among participants symptomatic at baseline (n = 535), the probability of remaining symptomatic declined steeply to 40.2% at the first follow-up, 19.3% at the second, and 7.6% by the last visit. In contrast, symptom absence was more stable: among those asymptomatic at baseline (n = 1,749), 81.1% remained asymptomatic at the first follow-up and 43.8% were still asymptomatic at the last visit (Supplementary Figure 1).

Symptom-level transitions revealed that cough (with or without sputum production) was the most frequently reported symptom among those developing symptoms (66%) and among those resolving symptoms (68%). Other common symptoms associated with symptom changes included chest pain, breathing difficulty, weight loss, and fever (Supplementary Figure 2).

When evaluating predictors for symptom transitions longitudinally among previously asymptomatic individuals, the onset of any TB symptom within the subsequent four-month interval was associated with a previous TB diagnosis (aRR 1.26; 95% CI 1.08-1.47; p=0.004), current smoking (aRR 1.81; 95% CI 1.53-2.15; p<0.001), past smoking (aRR 1.34; 95% CI 1.09-1.66; p=0.005), any drug use (aRR 1.23; 95% CI 1.08-1.41; p=0.002), and exposure to symptomatic cellmates (aRR 1.30; 95% CI 1.15-1.47; p<0.001). Among previously symptomatic individuals, resolution of symptoms was less likely among those who reported current smoking (aRR 0.82; 95% CI 0.72-0.92; p=0.001) or any drug use (aRR 0.88; 95% CI 0.79-0.97; p=0.010) (Supplementary Table 1). In interaction models, these predictors did not significantly modify the association between symptom transitions and TB risk.

Longitudinal analyses showed that compared to persistently asymptomatic individuals, transitioning from an asymptomatic to a symptomatic state within a four-month interval was significantly associated with incident TB (aRR 2.31; 95% CI 1.55-3.43). In contrast, remaining symptomatic (aRR 1.27; 95% CI 0.72-2.24) or resolution from symptomatic to asymptomatic (aRR 1.13; 95% CI 0.68-1.90) were not significantly associated with TB risk, compared to those who stayed persistently asymptomatic (Figure 1).

**Figure 1.**
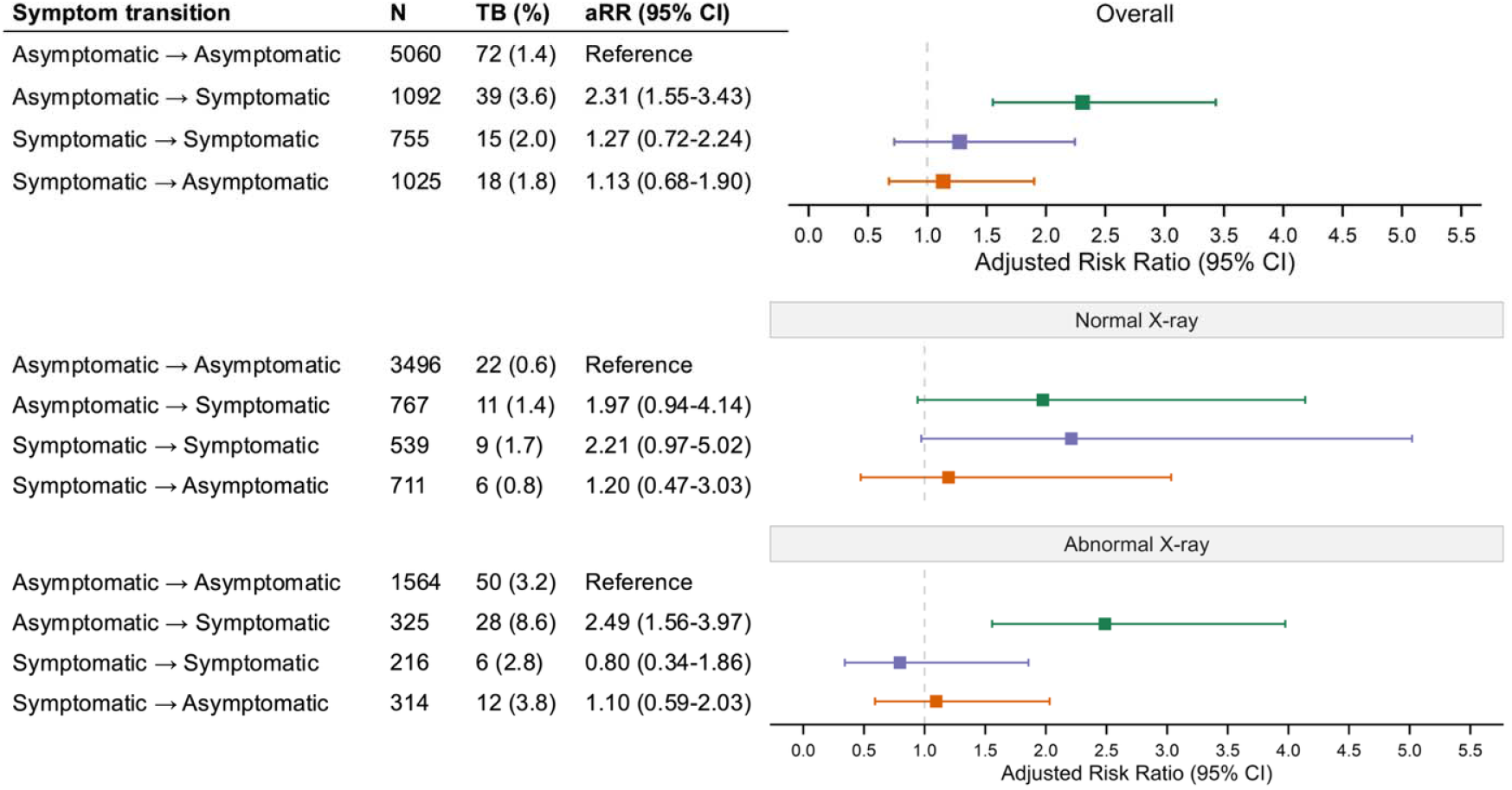
Adjusted risk of incident TB by symptom transitions over four-month intervals, overall and stratified by radiographic status N represents the total number of four-month symptom transition observations across all follow-up intervals during the study period. Adjusted risk ratios (aRRs) represent adjusted risk of TB diagnosed by sputum culture or Xpert within the same four-month interval as the symptom transition. Models adjust for baseline characteristics including age, prison, prior TB, smoking, drug use, duration of incarceration, symptomatic cellmates, and TB contact. The overall model adjusts for Lunit x-ray score category at the first visit of each interval. The stratified models adjust for the baseline characteristics and are separated by Lunit x-ray score category at the first visit of each interval. **Alt text:** Forest plot showing adjusted risk of incident tuberculosis by symptom transitions over four-month intervals. Compared with participants who remained asymptomatic, those who became symptomatic had over twice the risk of TB. Risks for persistent or resolving symptoms were similar to the reference group.

Stratified analyses revealed that this overall elevated risk among those developing symptoms was primarily driven by individuals with abnormal chest radiographs (aRR 2.49; 95% CI 1.56-3.97), though a similar trend was observed among those with normal x-rays (aRR 1.97; 95% CI 0.94-4.14). These findings indicate that symptom emergence was associated with elevated TB risk, with the strongest effects observed among those with radiographic abnormalities.

## DISCUSSION

In our longitudinal cohort study in two high-incidence prisons, we found that TB-related symptoms followed dynamic trajectories over four-month intervals. Between enrollment and the first follow-up visit just four months later, nearly one in seven (14.5%) developed new symptoms, and 60% of those symptomatic at baseline reported resolution, underscoring the fluctuating nature of TB-related symptom reporting in this population.

Importantly, more than half of all incident TB cases (52%, 26/50) occurred among participants who remained asymptomatic during the first interval, highlighting the substantial limitations of symptom-based screening in this setting. Prior studies in prisons and other high-risk congregate settings have similarly documented that reliance on cross-sectional symptom screening alone misses a considerable fraction of active TB cases, supporting the need for repeated active case-finding^4,14^. Our intensive screening program combining sputum testing and radiography every four months successfully detected microbiologically confirmed asymptomatic cases. Although our study was not designed to assess the additional yield of repeated symptom assessments, longitudinal monitoring may provide value in understanding disease progression, particularly when combined with complementary strategies such as routine sputum testing or radiographic evaluation.

Specifically, our analyses demonstrate that participants who developed new symptoms had more than double the risk of TB diagnosis compared to those who remained asymptomatic, while persistent symptoms conferred no excess risk. This pattern suggests that symptom emergence, rather than chronic symptom presence, offers greater resolution for identifying short-term risk. This may reflect the high sensitivity of our combined diagnostic strategy, which likely detected new TB cases rapidly, leaving few recent-onset cases undiagnosed. In contrast, participants with persistent symptoms may have alternative or unresolved etiologies, such as concomitant respiratory infections, which could obscure the relationship with incident TB. The overall association has practical implications for resource-limited settings where radiography may be unavailable, while stratified analyses show that symptom transitions most strongly predict TB risk among those with radiographic abnormalities, enabling more precise targeting of diagnostic resources.

We also identified key factors associated with four-month symptom transitions. Among previously asymptomatic individuals, previous TB, current and past smoking, and exposure to symptomatic cellmates increased the risk of developing symptoms. In contrast, current smoking and drug use were associated with persistent symptoms. While these predictors shaped symptom trajectories, they did not significantly modify the primary association between transitions and TB risk in interaction models. Nonetheless, combining repeated symptom assessments with knowledge of exposure-related risk factors may allow public health programs to more precisely target diagnostic interventions toward those at highest short-term TB risk.

Our study had several limitations. Symptom reporting relied entirely on self-report, which may be influenced by recall bias and subjective differences in symptom perception. Individuals who typically feel well may be more attuned to new symptoms, while those with chronic symptoms may have reduced sensitivity to incremental changes. However, our longitudinal assessment of symptom changes uses each individual as their own reference, potentially reducing the contextual and interviewer variability inherent in cross-sectional symptom reporting^15^. Additionally, the four-month interval between screenings may not have captured shorter-term or transient symptom fluctuations. Excluding participants with intermittently missing visits may have introduced bias, as those excluded were more likely to report drug use and lower education, and may have had less stable follow-up due to transfer or refusal. We also lacked reliable data on nutrition and longitudinal smoking behavior, precluding assessment of these important TB risk factors. Despite these limitations, our findings offer important insights into the temporal dynamics of TB symptoms and their potential utility in prioritizing individuals at highest risk for diagnostic screening and treatment.

In conclusion, longitudinal assessment of TB-related symptoms reveals dynamic patterns of emergence and resolution that identify individuals at elevated short-term risk of active disease. Algorithms prioritizing individuals with either new-onset symptoms or radiographic abnormalities for molecular testing may improve sensitivity and cost-effectiveness in high-transmission, resource-limited settings. A recent study found that symptom screening, while less sensitive than annual radiography, can achieve comparable impact when applied frequently and at lower cost^16^. Our findings suggest that monitoring symptom transitions may enhance this existing approach. Future studies should assess how integrating longitudinal symptom monitoring, radiography, and rapid diagnostics can be optimized to reduce transmission across high-burden settings.

## Data Availability

Analytic code is available at https://github.com/Andrews-Lab-Stanford/LongitudinalTBSymptoms_BrazilPrisons.git

## Acknowledgements

EJ, JRA, and JC conceived of the study. JRA, JC, and RDO developed the study plans. AmdS and AdSS enrolled participants and collected data. DHT performed data management. EJ, JVBB, YL, ADS, and JRA designed and performed the data analysis. EJ and JVBB wrote the manuscript. All authors contributed to revising the manuscript.

The authors acknowledge the use of ChatGPT (GPT-4, OpenAI) for troubleshooting coding errors and minor grammar checking. AI was not used for generation of conclusions or interpretation of results. All content and analyses were independently verified by the authors, who take full responsibility for the manuscript.

## APPENDIX

**Supplementary Figure 1.**
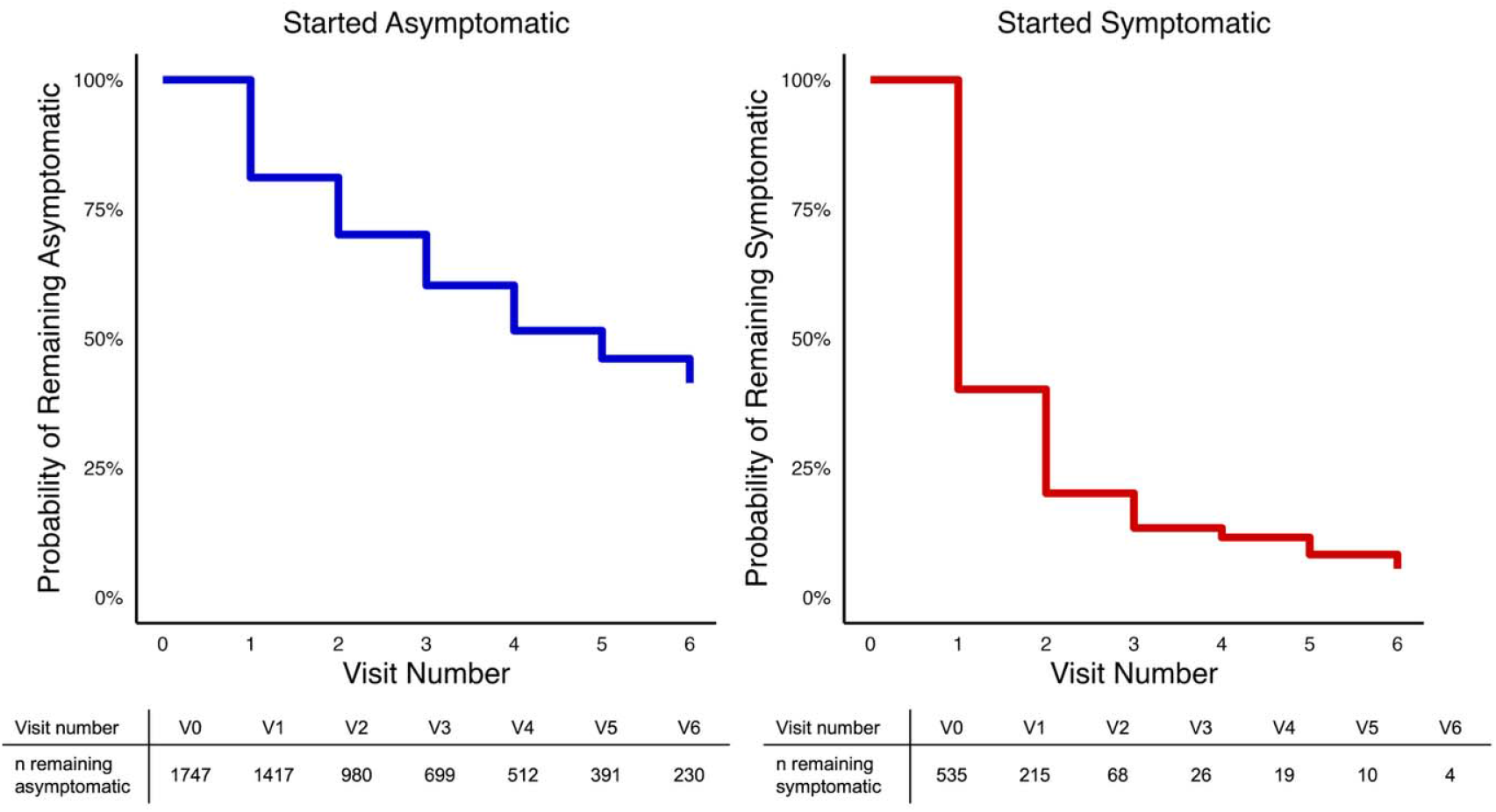
Symptom persistence by baseline symptom status Risk tables with raw numbers of participants at each visit are shown beneath the curves. **Alt text:** Kaplan-Meier curves showing symptom persistence by baseline symptom status. Participants starting asymptomatic had a gradual decline in remaining asymptomatic, while those starting symptomatic showed rapid symptom resolution over follow-up visits.

**Supplementary Figure 2.**
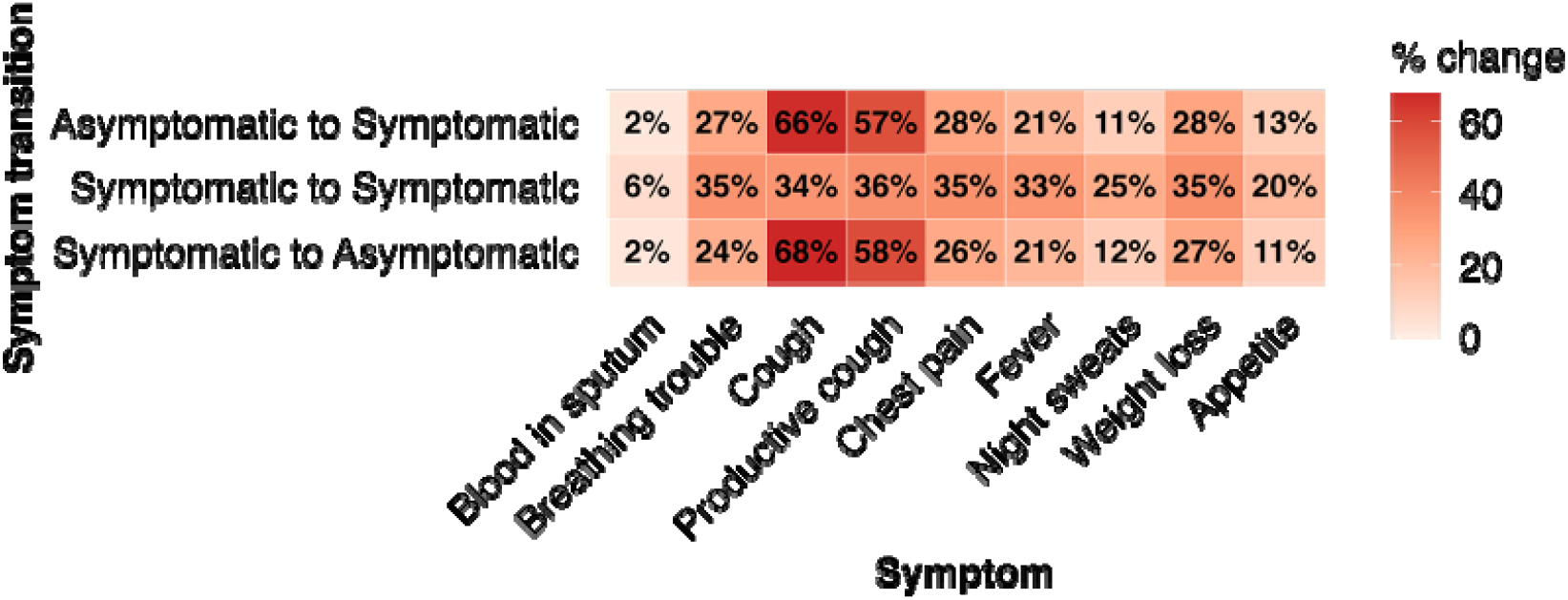
Specific symptom changes by symptom transition group % of total instances of symptom transition in that symptom category. Symptom changes refer to new onset (Asymptomatic to Symptomatic group), resolution (Symptomatic to Asymptomatic), or either (Symptomatic to Symptomatic). **Alt text:** Heatmap showing percentage changes in specific symptoms by symptom transition group. Color intensity reflects percent change, highest among transitions involving cough-related symptoms.

**Supplementary Figure 3.**
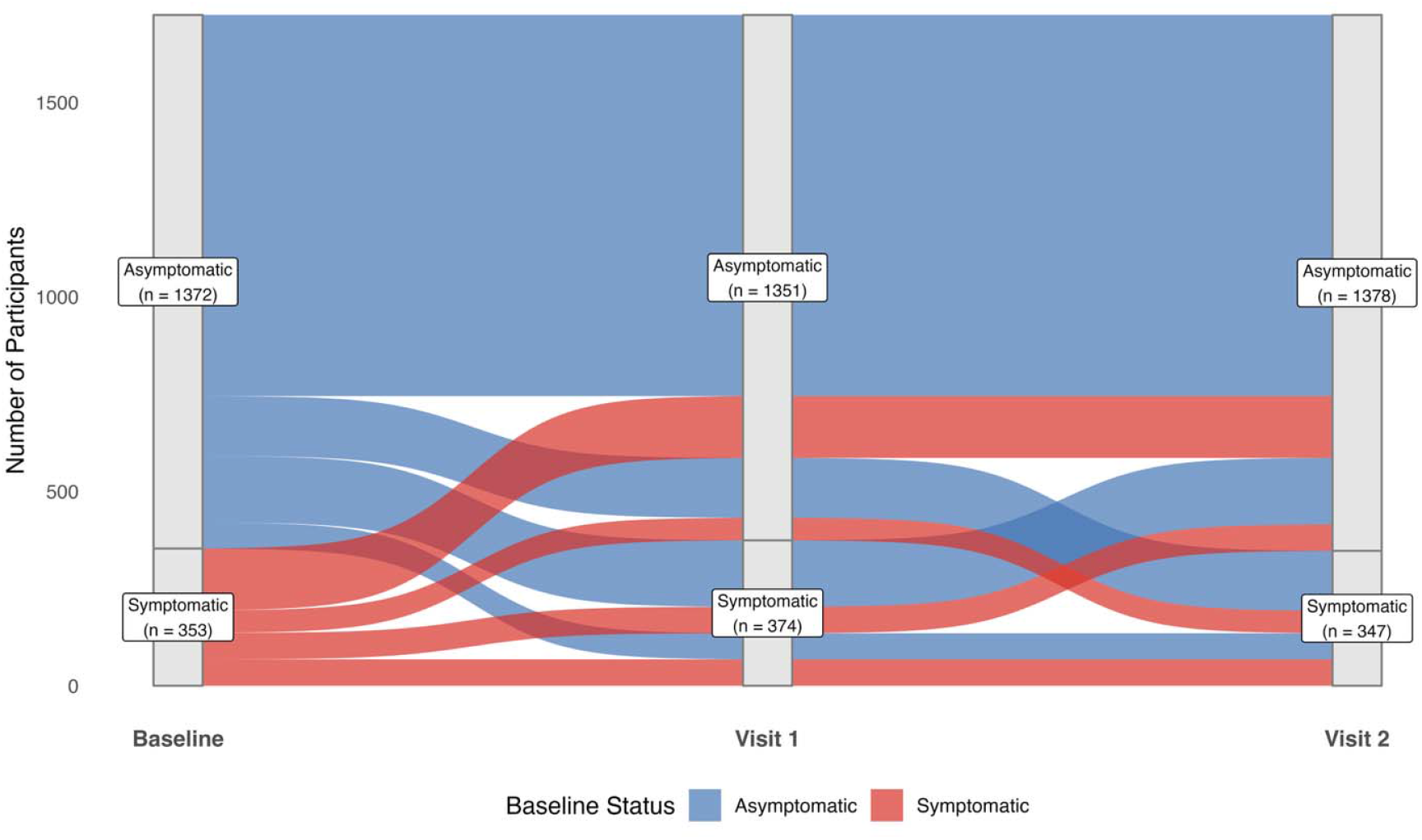
Symptom transitions among participants with at least three symptom assessments (N = 1725) **Alt text:** A Sankey diagram visualizing symptom state transitions for 1,725 participants across three timepoints: Baseline, Visit 1, and Visit 2. The chart illustrates frequent bidirectional movement between asymptomatic and symptomatic states, highlighting the dynamic nature of symptoms over time.

**Supplementary Table 1.** Baseline predictors of symptom change stratified by prior symptom status.

| Baseline predictors | Becoming symptomatic |  |  |
| --- | --- | --- | --- |
|  | aRR* | 95% CI | p-value |
| Previous TB | 1.26 | (1.08-1.47) | 0.004 |
| Current smoking | 1.81 | (1.53-2.15) | <0.001 |
| Past smoking | 1.34 | (1.09-1.66) | 0.005 |
| Any drug use | 1.23 | (1.08-1.41) | 0.002 |
| Symptomatic cellmates | 1.30 | (1.15-1.47) | <0.001 |
| TB contact | 1.03 | (0.91-1.17) | 0.662 |
| LunitTB ≥50 | 0.90 | (0.78-1.04) | 0.164 |

Previously symptomatic
| Baseline predictors | Becoming asymptomatic |  |  |
| --- | --- | --- | --- |
|  | aRR* | 95% CI | p-value |
| Previous TB | 0.94 | (0.83-1.06) | 0.340 |
| Current smoking | 0.82 | (0.72-0.92) | 0.001 |
| Past smoking | 0.96 | (0.83-1.10) | 0.563 |
| Any drug use | 0.88 | (0.79-0.97) | 0.010 |
| Symptomatic cellmates | 0.92 | (0.84-1.00) | 0.057 |
| TB contact | 0.92 | (0.84-1.01) | 0.077 |
| LunitTB ≥50 | 0.98 | (0.88-1.10) | 0.784 |

\*Adjusted risk ratios (aRRs) represent risk of symptom status change from prior status compared to no change in symptom status. Multivariate logistic regression models with cluster-robust standard errors at the individual level to estimate odds of symptom transitions, adjusting for baseline characteristics including age, prison, prior TB, smoking, drug use, prison time, symptomatic cellmates, TB contact, and categorical LunitTB x-ray score.

